# Analysis of the increase in cross-sectional area of the median nerve and its relation to age in neonates, infants and children using high-resolution ultrasound imaging

**DOI:** 10.1101/2020.05.22.20110080

**Authors:** Carole Jenny, Jürg Lütschg, Philip J. Broser

**Author notes:** Correspondence to: Dr. Philip J. Broser, Ostschweizer Kinderspital, Neuropediatric Department, Claudiusstrasse. 6, CH – 9006 St. Gallen.

## Abstract

**AIM:** To analyze the increase in the cross-sectional area (CSA) of the median nerve during early childhood.

**METHOD:** This prospective, cross-sectional study used high-resolution ultrasound images of the median nerve from three locations (wrist, forearm and upper arm). A total of 43 participants (32 of whom were children younger than 2 years) were included in the study.

**RESULTS:** A strong and highly significant correlation was found between age and CSA (r = 0.8, *p* < 0.0001). The growth rate of CSA decreases with age. The increase in CSA follows a logarithmic growth curve (*p* < 0.0001). Based on the regression analysis, an age-synchronous increase in CSA for all three locations was found. The nerve reaches 70% of its final CSA by 2 years of age.

**INTERPRETATION:** Similar to the nerve conduction speed, the increase in CSA is greatest during the first 2 years of life. Then, the rate gradually and synchronously slows at the proximal and distal locations.

**What this paper adds:**

- Normative values for increased cross-sectional area (CSA) of the median nerve.
- Standardized locations and image procedures outlined for the clinical setting.
- Growth dynamic of the CSA of the median nerve in children.
- Normative data for development of the median nerve in children.
- High resolution ultrasound images of the maturating median nerve in neonates, infants and children.

---

During the first years of life, rapid development of many psychomotor functions occurs. One important part of the development of the central and peripheral nervous system^1^ is an acceleration of the axonal transmission of information. Raimbault^2^ showed that the motor and sensory nerve conduction velocity almost doubles during the first 2 years of life. This increase in nerve conduction velocity is due to a structural change in the peripheral nerves, especially an increase in myelination. This increase in myelination leads to an increase in the cross-sectional area (CSA) of the nerves.^3^

Several studies have analyzed the size of peripheral nerves in children using ultrasound imaging.^4–7^ However, most of these studies focused on children over 2 years of age.^4^ Until recently, it has been difficult to image and precisely measure even the major nerves in neonates. This is because at the time of birth, even the median nerve has a CSA of only 1.5 mm^2^. Therefore, little is known about how the size of the nerve’s changes during the critical developmental period from birth to 2 years. It recently became possible to image and quantify the peripheral nerves in children and newborns using high-frequency ultrasound devices precisely.

The aim of this study is to analyze the age dependency of the increase in the CSA of the median nerve at three locations (wrist, forearm and upper arm; Figure 1A, B) during the first 2 years of life (main study group). In order to compare the results of this study with previously published data, a select number of children older than 2 years (comparison group) were also included.

**Figure 1:**
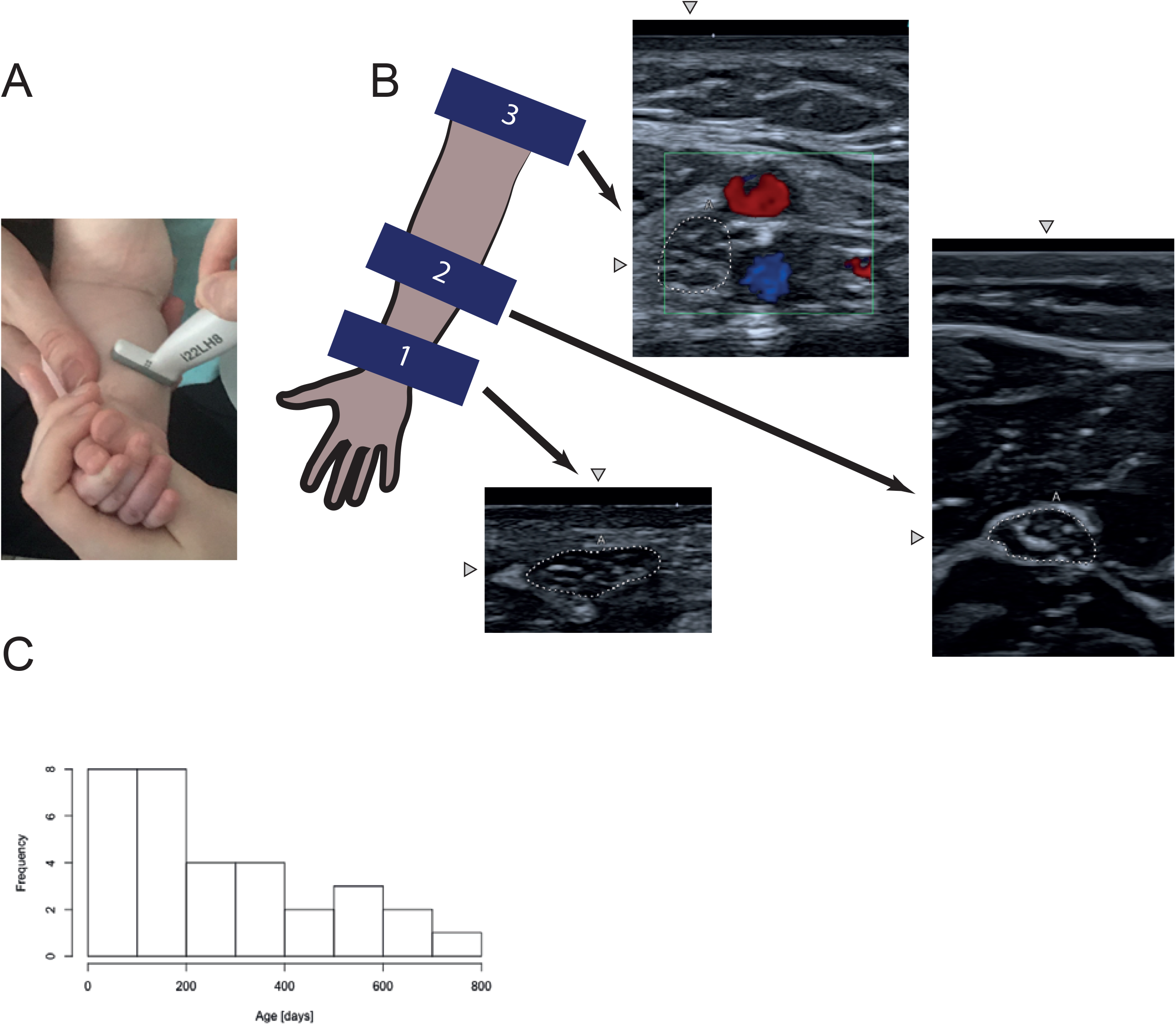
(A) Placement of ultrasound probe (Canon i22LH8) on the forearm to image the median nerve at location 2. Child’s image used with permission. (B, left) Positioning of the three imaging locations schematically on the right arm. (B, right) Sample images for Locations 1–3. White dotted line is the median nerve. Arrowheads point to the median nerve. (C) Age distribution of the subjects in the main study group (age < 2 years). Each age segment is 100 days.

## METHOD

This prospective cross-sectional study was conducted at the children’s hospital of Eastern Switzerland from December 2019 to March 2020. The study was approved by the local ethical committee and registered with the Swiss project database (RAPS, EKOS Nr. 19/166). Written informed consent was obtained from the caregivers prior to inclusion of participants in the study.

Typically developing children between 0 days and 10 years were eligible for inclusion in the study. Children below the age of 2 years were assigned to the main study group; all older children were assigned to the comparison group. Children with a chronic disease, premature birth, severe acute disease or family history of any inherited neurological disease were excluded from the study. Children were solely recruited at the children’s hospital of Eastern Switzerland. The children were either hospitalized for mild non-chronic diseases (e.g., human respiratory syncytial virus infections) or were siblings of patients. Imaging was typically performed just before discharge from the hospital. The authors planned a priori to recruit a total of 50 patients, with at least 30 participants in the main study group. The age range of the main study group was further divided into intervals of 100 days, and patients were screened with the aim of recruiting at least three participants for each interval. Apart from the screening procedure, there was no difference between the main group and comparison group.

A Canon Aplio i800 (Canon Medical Systems, Tokyo, Japan) ultrasound imaging system equipped with i33LX9 (max scanning frequency of 33 MHz) and i22LH8 (max scanning frequency of 22 MHz) ultrasound probes was used for imaging. The ‘penetration’ setting, with an imaging depth of 15 mm, was used for the i33LX9 probe (footprint of 35 mm), and the ‘general’ setting, with an imaging depth of 17.5 mm, was used for the i22LH8 probe (footprint of 26 mm). This gave an axial resolution in the range of 30 μm for the i33LX9 probe and 50 μm for the i22LH8 probe.

For each participant, the median nerve was imaged at three locations (Figure 1A, B).^8,9^ Location 1, the wrist, was scanned at the level of the wrist (close to the musculus pronator quadratus). Location 2, the mid-forearm, was scanned halfway between wrist and elbow. At this location, the median nerve is located above the deep flexor muscles and below the superficial flexors. Location 3, the upper arm, was scanned proximal of the elbow (lower third of the upper arm). Here, the median nerve is located just adjacent to the brachial artery. Therefore, for this location, ultrasound doppler was used to localize the brachial artery in order to identify the nerve. Both arms were examined if possible. However, this was not always possible due to medical equipment attached to the patient (e.g., intravenous cannulas) or a lack of compliance.

For all three locations, transverse orthogonal still images of the median nerve were recorded. The CSA and circumference were measured from the stored still images after the examination to reduce the time required for the child to hold still. Given that ultrasound images were taken systematically at the same locations, the nerve was easily identified in these images (Figure 1B).

After identifying the nerve on the image, the ultrasound reflection border between connecting tissue and the nerve was manually traced in the ultrasound system using the freehand tracing tool (Figure 1B). After manual tracing, the tool provided the CSA in square millimetres and circumference of the nerve in millimetres. Both values were stored for all three locations. If available, the age, sex, height and weight of the patients were recorded and stored with the imaging data in the study database.

To reduce inter-observer variability, the imaging and measurement procedure and personnel were kept strictly the same for all participants. The participants were recruited and prepared for scanning by author CJ. The imaging and tracing was also performed by CJ under the supervision of author PB.

All statistical computation was conducted using the R statistic program.^10^ Two sets of scatter plots relating the age of the participants to the CSA of the nerve were generated. The first sets included both the main study group and comparison group. The second set focused specifically on the main study group.

A two-sided Student’s t-test was used to test if the CSA values on the left and right side were significantly different. Age-matched pairs of female and male participants were built to test differences in relation to sex using Student’s t-test.

The correlation between age, weight and CSA was tested for statistical significance using Pearson’s correlation.^11^ Logarithmic regression analysis was performed to determine if a logarithmic model could describe the relationship between age and CSA.^12^ Regression analysis was performed on the data from the right median nerve. The critical p-value was set to 5 % and if necessary was corrected for multiple comparisons using Bonferoni’s method.^13^

## RESULTS

A total of 84 patients were screened to obtain the target number of 50 patients. Seven participants had to be removed from the study (three due to missing informed consent, two due to images not properly stored and two due to caregivers withdrawing from the study).

A total of 43 children (19 males) between the ages of 12 days and 10 years were included in the final study. Of these, 32 children were younger than 2 years and were thus included in the main study group. The distribution of participants in the main study group is shown in Figure 1C. For two age ranges (400–500 days and 600–700 days), only two participants could be recruited, and the for the age range 700–800 days, only one participant was recruited. The complete list of demographic data for the participants is shown in Table I, left column.

**Table I:** Demographics and median nerve measurements. Area = cross-sectional area, Circ. = circumference. IDs were assigned by the ultrasound machine and are not necessarily consecutive. (Remark: To optimize the layout of the table. The table has been uploaded as a PDF document.)

Median nerve area and circumference as measured by high resolution ultrasound imaging
| ID | Age(days) | Weight(KG) | Size (cm) | Gender | Median nerve area and circumference as measured by high resolution ultrasound imaging |  |  |  |  |  |  |  |  |  |  |  |
| --- | --- | --- | --- | --- | --- | --- | --- | --- | --- | --- | --- | --- | --- | --- | --- | --- |
|  |  |  |  |  | Right side |  |  |  |  |  | Left side |  |  |  |  |  |
|  |  |  |  |  | Location 1 |  | Location 2 |  | Location 3 |  | Location 1 |  | Location 2 |  | Location 3 |  |
|  |  |  |  |  | Area R1 | Circ. R1 | Area R2 | Circ. R2 | Area R3 | Circ.R3 | Area L1 | Circ. L1 | Area L2 | Circ. L2 | Area L3 | Circ. L3 |
| 9 | 12 | 2.82 | 50.50 | f | 2.01 | 6.72 | 1.49 | 4.66 | 1.52 | 5.84 | 1.91 | 7.27 | 1.85 | 5.37 | 1.58 | 5.41 |
| 7 | 18 | 3.98 | 51.10 | f | 1.62 | 7.44 | 1.00 | 3.82 |  |  | 2.97 | 8.67 | 2.21 | 5.65 |  |  |
| 58 | 24 | 4.54 | NN | f | 2.19 | 8.54 | 1.89 | 5.22 | 3.04 | 6.44 | 2.28 | 6.38 |  |  |  |  |
| 56 | 25 | 4.20 | 51.50 | m | 1.73 | 7.20 | 1.60 | 4.72 | 1.75 | 4.92 | 1.71 | 6.57 | 1.53 | 4.63 | 1.77 | 5.08 |
| 51 | 26 | 3.39 | 50.50 | f | 2.12 | 7.24 | 1.49 | 4.57 | 2.57 | 7.13 | 1.69 | 6.44 | 2.04 | 5.71 | 2.50 | 5.91 |
| 47 | 52 | 6.18 | 60.10 | m | 3.25 | 7.48 | 2.52 | 6.07 | 3.18 | 7.52 |  |  |  |  |  |  |
| 4 | 68 | 5.07 | 55.6 | m | 2.10 | 6.75 | 1.96 | 5.84 |  |  | 2.07 | 8.45 |  |  |  |  |
| 31 | 70 | 5.46 | 56.80 | f | 2.80 | 7.82 | 1.60 | 6.06 | 3.19 | 7.24 | 2.82 | 7.83 | 1.53 | 4.57 | 3.27 | 8.51 |
| 17 | 107 | 6.07 | 61.00 | m | 3.93 | 8.39 | 1.69 | 5.07 | 3.14 | 7.05 |  |  |  |  |  |  |
| 48 | 146 | 6.87 | 65.00 | f |  |  |  |  |  |  | 2.40 | 7.37 | 2.12 | 5.66 | 2.82 | 7.26 |
| 10 | 159 | 8.20 | NN | m | 2.42 | 8.11 | 1.78 | 5.92 | 2.80 | 6.44 | 1.21 | 6.40 | 2.35 | 6.67 | 2.39 | 6.38 |
| 30 | 159 | NN | NN | f | 2.62 | 7.76 | 2.67 | 5.96 | 3.57 | 7.82 | 3.14 | 7.52 | 2.35 | 6.53 | 3.68 | 7.40 |
| 34 | 176 | 6.59 | NN | f | 2.31 | 7.83 | 2.72 | 6.46 | 3.90 | 7.97 |  |  |  |  |  |  |
| 5 | 179 | 7.84 | 70.00 | m | 2.60 | 9.12 |  |  |  |  | 2.45 | 6.46 |  |  |  |  |
| 22 | 180 | 8.17 | NN | f | 2.51 | 6.75 | 2.36 | 6.49 | 3.49 | 7.02 | 3.39 | 7.44 | 1.64 | 5.93 | 3.00 | 7.09 |
| 52 | 193 | 7.59 | 66.00 | m | 3.17 | 8.26 | 2.71 | 6.17 | 3.22 | 7.29 | 3.22 | 8.30 | 2.91 | 6.58 | 3.56 | 8.12 |
| 29 | 220 | 8.04 | NN | m | 3.65 | 8.00 | 2.88 | 6.73 | 3.90 | 7.58 | 3.62 | 7.79 | 2.31 | 6.35 | 4.60 | 8.38 |
| 18 | 248 | 8.10 | 73.00 | f | 2.95 | 9.25 | 3.18 | 6.67 |  |  |  |  |  |  |  |  |
| 35 | 248 | 6.79 | 71.00 | f | 2.99 | 7.96 | 1.84 | 5.08 | 3.60 | 8.43 | 3.12 | 8.30 | 2.13 | 5.89 | 3.09 | 6.88 |
| 36 | 279 | 8.5 | NN | m | 3.15 | 9.36 | 2.09 | 5.88 | 3.20 | 7.13 |  |  |  |  |  |  |
| 14 | 362 | 11.00 | 78.50 | f | 3.34 | 8.90 | 1.60 | 5.73 | 3.92 | 8.53 |  |  |  |  |  |  |
| 32 | 387 | 9.70 | 78.00 | f | 3.16 | 8.73 | 2.63 | 6.78 | 4.41 | 7.84 | 2.92 | 7.90 | 1.90 | 5.17 | 3.63 | 7.70 |
| 57 | 387 | 7.08 | 71.00 | m | 3.12 | 8.22 | 2.27 | 5.88 | 3.86 | 8.09 | 2.87 | 8.38 | 1.91 | 5.41 | 3.81 | 8.52 |
| 21 | 389 | 10.60 | 74.00 | f |  |  | 2.89 | 6.53 |  |  |  |  |  |  |  |  |
| 26 | 464 | 9.20 | 76.00 | f | 3.88 | 10.71 | 2.43 | 6.61 | 4.43 | 8.67 | 4.61 | 10.66 | 2.81 | 6.20 | 3.57 | 7.02 |
| 44 | 464 | 10.20 | 77.00 | w | 3.88 | 9.95 | 3.11 | 7.19 | 4.31 | 8.36 | 3.70 | 10.63 | 2.79 | 6.85 | 4.18 | 8.44 |
| 33 | 502 | NN | NN | f | 3.83 | 8.59 | 3.17 | 8.28 | 4.30 | 8.83 | 3.59 | 10.32 | 2.11 | 5.84 | 4.78 | 9.96 |
| 8 | 512 | 10.15 | NN | f | 3.53 | 9.47 | 2.71 | 6.92 |  |  | 3.40 | 9.07 | 3.92 | 7.83 |  |  |
| 50 | 590 | 11.00 | NN | m | 3.77 | 7.53 | 3.47 | 7.26 | 4.28 | 8.27 | 3.60 | 11.30 | 3.40 | 6.87 | 4.08 | 7.81 |
| 15 | 659 | 11.00 | 89.00 | f | 5.09 | 9.41 | 3.09 | 6.84 | 4.43 | 7.98 | 4.45 | 9.52 | 2.40 | 5.71 | 3.09 | 7.10 |
| 49 | 670 | 10.70 | 88.70 | f | 3.37 | 8.89 | 2.38 | 6.13 | 4.00 | 8.48 | 3.39 | 7.97 | 2.41 | 6.39 | 3.49 | 7.94 |
| 6 | 720 | 15.00 | NN | m |  |  | 2.98 | 6.97 | 6.12 | 9.36 | 5.46 | 12.26 | 2.77 | 6.99 |  |  |
| 45 | 748 | 10.50 | 85.00 | f | 4.26 | 9.13 | 2.68 | 6.40 | 4.78 | 8.76 | 4.29 | 9.65 | 3.25 | 7.37 | 4.68 | 8.04 |
| 43 | 775 | 15.40 | 94.50 | m | 4.94 | 11.13 | 3.68 | 7.33 | 4.82 | 8.92 | 4.26 | 8.25 | 2.71 | 6.14 | 3.69 | 7.38 |
| 37 | 1093 | 13.40 | 94.60 | m | 3.52 | 10.00 | 2.97 | 6.44 | 4.09 | 8.45 | 3.85 | 9.32 | 2.21 | 5.61 | 5.29 | 8.64 |
| 46 | 1532 | 20.1 | NN | m | 4.38 | 9.42 | 4.22 | 8.45 | 4.44 | 7.71 | 4.34 | 10.21 | 4.24 | 7.79 | 5.38 | 9.23 |
| 61 | 1600 | NN | NN | m | 4.79 | 11.25 | 3.37 | 7.03 | 7.48 | 11.04 | 4.9 | 10.36 | 3.14 | 6.96 | 5.38 | 9.2 |
| 62 | 1827 | 16.8 | 104 | f | 3.71 | 10.26 | 3.05 | 6.95 | 4.18 | 8.07 | 3.72 | 9.77 | 3.39 | 7.28 | 3.01 | 7.95 |
| 59 | 2200 | 19.70 | NN | f | 4.16 | 10.54 | 3.78 | 7.29 | 4.74 | 8.64 |  |  |  |  |  |  |
| 60 | 2211 | 20.80 | 119.5 | m | 4.72 | 9.99 | 3.59 | 7.52 | 4.91 | 8.04 | 5.16 | 9.51 | 4.03 | 7.79 | 5.62 | 11.54 |
| 55 | 3341 | NN | NN | m | 6.32 | 11.11 | 5.04 | 8.76 | 5.30 | 8.42 | 6.08 | 10.43 | 4.15 | 7.84 | 5.82 | 8.84 |
| 53 | 3454 | 27.00 | NN | f | 5.89 | 11.56 | 3.21 | 6.60 | 7.58 | 10.47 |  |  |  |  |  |  |
| 54 | 4007 | 33.00 | 151.00 | m | 6.27 | 12.35 | 4.31 | 7.89 | 9.09 | 13.08 |  |  |  |  |  |  |

### Size of the cross-sectional area

Figure 2 shows examples of recorded still images from five participants in order by ascending age. These and similar images were used to measure CSA for each participant.

**Figure 2:**
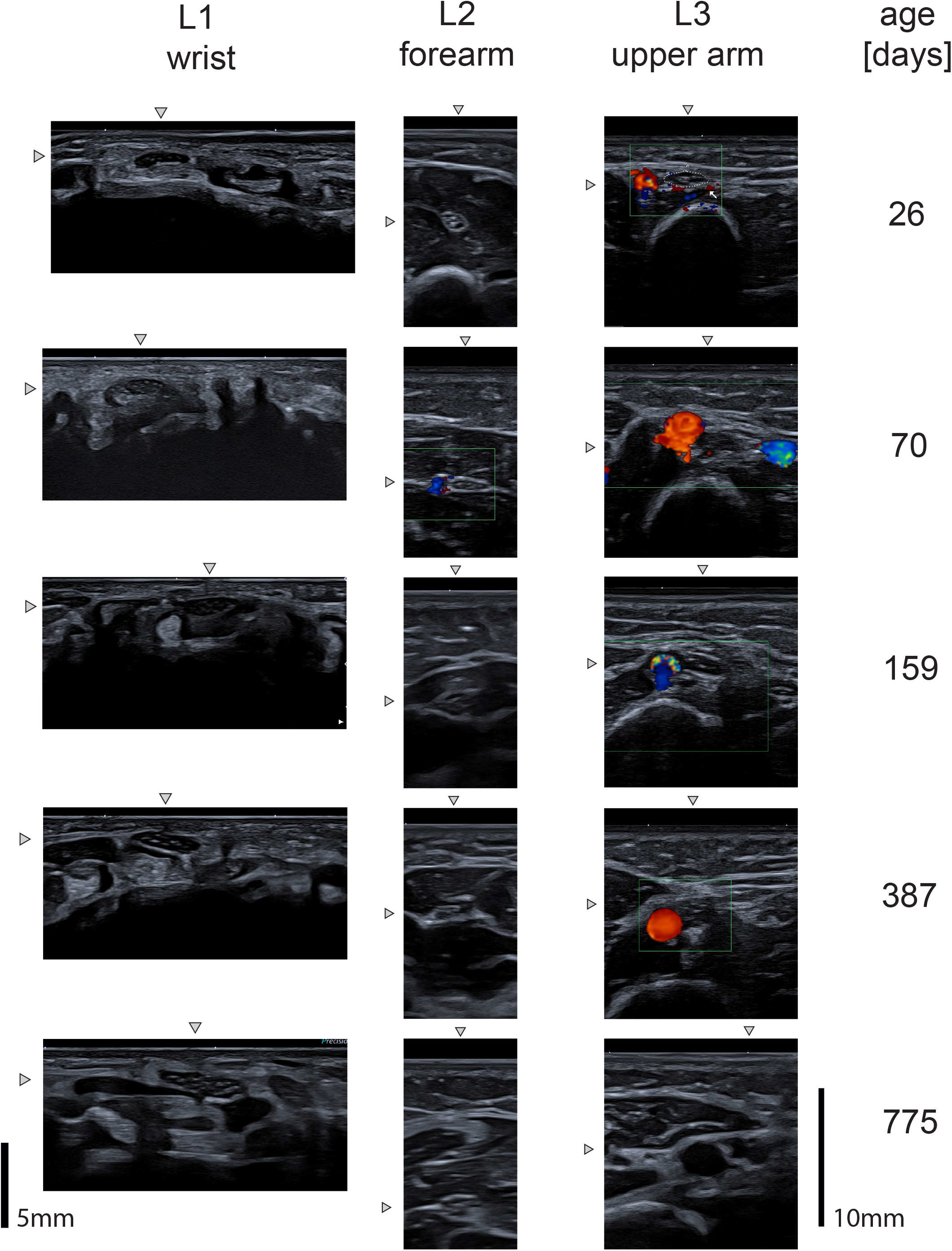
Typical ultrasound images for all three locations from five participants ages 26 days to 775 days. Arrowheads point in the direction of the nerve. The nerve is located at the intersection of the two directions. The scale bar on the lower left applies to all images of Location 1, and the scale bar on the lower right applies to all images of Locations 2 and 3.

The measurements are listed in Table I, right column. For those participants for whom the measurement data for the left and right median nerve were available, the CSA values were compared using Student’s paired t-test to determine the difference. No significant difference was found between the left and right median nerves for Location 1 (n = 31, mean difference R-L: 0.002 mm^2^, *p* > 0.05) or Location 2 (n = 29, mean difference R-L: 0.06 mm^2^, *p* > 0.05). A significant difference was found for Location 3 (n = 26, mean difference R-L: −1.05 mm^2^, *p* < 0.001). The age-matched pairs did not show any significant difference in CSA in relation to sex (Locations 1–3: mean difference m-f < 0.4 mm^2^, *p* > 0.1).

Visual inspection of the images revealed an increase in the size of the nerve with age (Figure 2). To quantify this qualitative finding, scatter plots relating age and CSA are shown in Figure 3. Further Pearson’s correlation tests were applied to test the correlation between age and CSA for the three locations. The results showed highly significant correlations for all three locations (Location 1: r = 0.83, *p* < 0.0001; Location 2: r = 0.75, *p* < 0.0001; Location 3: r = 0.80, *p* < 0.0001), confirming a statistically significant increase in CSA with age for all three locations.

**Figure 3:**
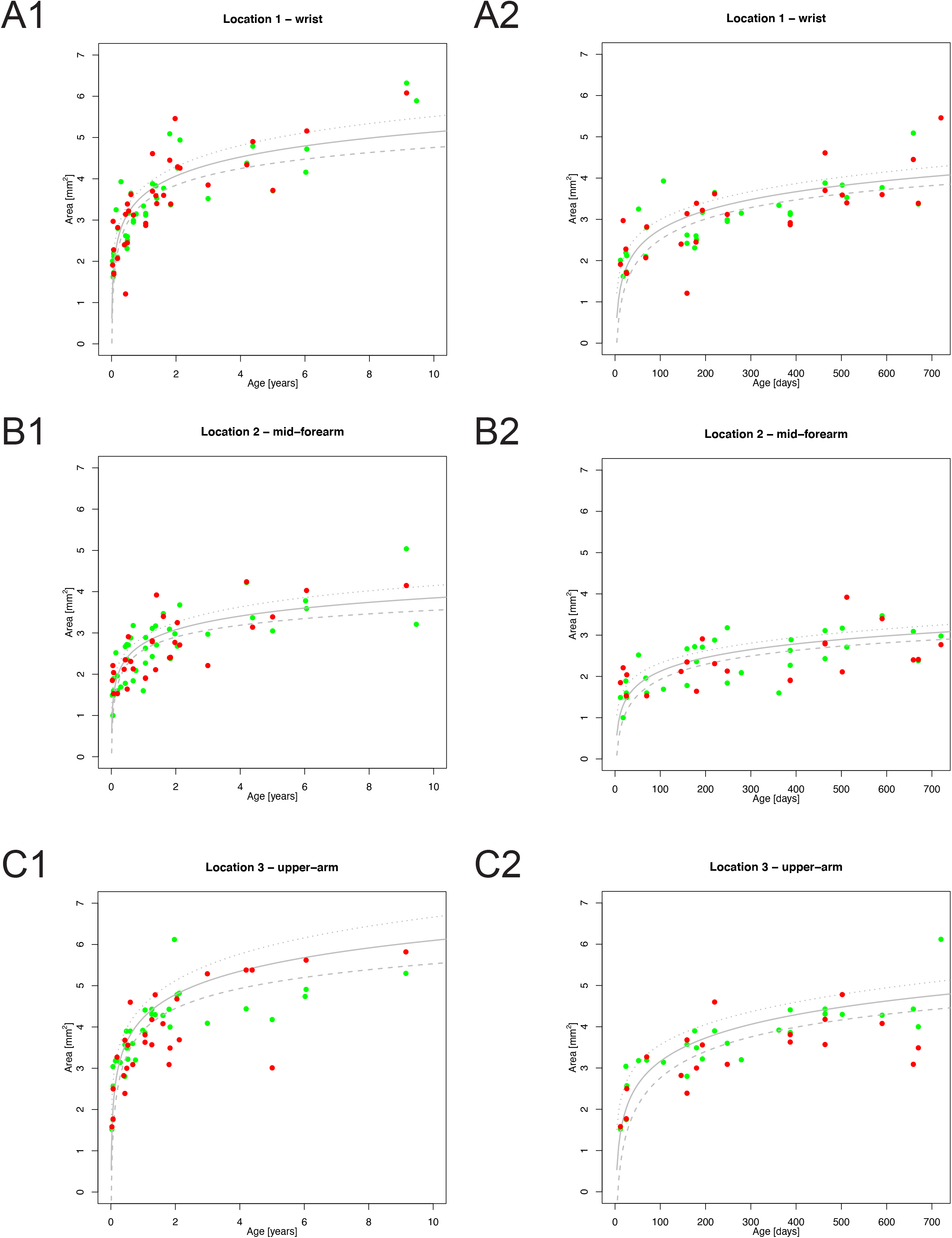
(A) Location 1: wrist; (B) Location 2: forearm; (C) Location 3: upper arm. The measured cross-sectional area is plotted against age (green dots: right arm; red dots: left arm). Panels A.1–C.1 cover the whole age range of the study (main study group and comparison group). Panels A.2–C.2 focus on the main study group. The logarithmic regression curve is plotted in each panel as a grey line with the limits of the lower and upper 95% confidence interval plotted as dotted lines above and below.

### Logarithmic regression curve

The scatter plots in Figure 3 show that the increase in CSA is most significant during the first 2 years of life and then gradually slows with increasing age. This is typical behaviour for processes that are governed by a logarithmic model. Therefore, the model:

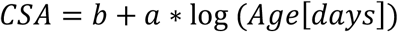

was chosen, and a regression analysis as described by Natale and Rajagopalan^14^ was performed *(a: slope* and *b: intersection)*.

For each location, the model was tested and *a* and *b* were numerically determined by the regression analysis (Location 1: *b* = −0.30, *a* = 0.66, *p* < 0.0001; Location 2: *b* = −0.08, *a* = 0.48, *p* < 0.0001; Location 3: *b* = −0.60, *a* = 0.82, *p* < 0.0001). In summary, the regression analysis showed a highly significant result, suggesting that the increase in CSA is well-described by the logarithmic model. The regression curve is shown in Figure 2A.1–C.1. According to these logarithmic models, the nerve reaches 70% of its approximated final CSA by the age of 2 years for all three locations.

It is well-documented that during the first years of life, children’s age and weight are closely correlated.^14^ As a consequence of this correlation and the correlation between age and CSA, a correlation between weight and CSA was expected. Therefore, a further correlation analysis was performed correlating weight and CSA. Pearson’s correlation test showed a highly significant correlation between the two (Location 1: r = 0.85, *p* < 0.0001; Location 2: r = 0.80, *p* < 0.0001; Location 3: r = 0.89, *p* < 0.0001). Supplementary Figure 1 shows scatter plots of the CSA in relation to weight and age.

### Discussion

In this study, the growth of the CSA of the median nerve during the first 2 years of life was analyzed at three peripheral locations. A highly significant correlation was found between age and CSA for all three locations. It was further shown that the growth of the CSA is governed by a logarithmic model. Given that age and weight are significantly correlated during early childhood,^14^ a further analysis correlating weight and CSA was conducted. As expected, this correlation was also highly significant. The sex of the participants had no measurable effect on the CSA. The difference in CSA between the left and right median nerve was also tested for significance. For the CSA at the wrist (Location 1) and mid-forearm (Location 2), no significant difference was found. However, at the upper arm (Location 3), the CSA was 20% smaller for the right side than the left. Similar size differences of up to 30% have been previously reported for children older than 2 years.^5^

One limitation of this study is that it only analyzed about three children per 100-day interval for the age range of 0 to 2 years. In addition, the study was designed as a cross-sectional study, but a longitudinal study design would probably be more appropriate to monitor the increase in CSA with age, especially during the first 2 years of life. Further, the analysis was restricted to the measurement of the median nerve at the arm. The development at more proximal locations, especially around the fascicle and next to the nerve roots, could differ. In addition, the growth dynamic of the lower extremities could also be different.

Based on the growth data in this study, some conclusions can be drawn that reveal characteristics of the maturation of the peripheral nervous system. First, the logarithmic model of the increase in CSA and its first derivative according to time shows that the growth rate of CSA is inversely proportional to age. This result suggests that the growth rate is highest at the time of birth and then gradually decreases with age. This is obvious for the data presented in this study and corresponds with the data for older children^5^ and even for adults.^10^ The correlation and normalization of the CSA to weight and age (Supplementary Figure 1) further shows that the CSA for the median nerve converges at a value of about 0.1 mm^2^/kg (bodyweight).

According to the work by Gesslbauer,^16^ the median nerve at the level of the upper arm consists of about 41,000 axons (about 1.3% of axons are from motor neurons). This number is relatively stable between individuals and stays almost constant throughout life.^16^ Dividing the CSA of the median nerve by the number of axons of the nerve estimates the average area that can be occupied by one axon and its myelin shed. Given that the nerve consists of unmyelinated, weak and strongly myelinated fibres, and given that the nerve has a fascicular structure with connecting tissue between the fascicle, this is only a gross estimate but still quite informative. This calculates to 2 mm^2^/40,000 axons = 50 μm^2^/axon at birth and 4 mm^2^/40,000 axons = 100 μm^2^/axon at 2 years of age. A medium-sized axon has a radius of about 5 μm and a myelin shed of about 2 μm and therefore typically occupies a space of 153 μm^2^. Thus, the increase in CSA gives each axon more space and therefore facilitates myelinization.

The study presented here further shows that for all three (peripheral) locations, the median nerve reaches 70% of its final CSA by approximately 2 years of age. This suggests that the nerve matures similarly at all three peripheral locations. This finding is in accordance with the histological data by Schröder et al.^4^ However, it is well-known that the motor roots maturate before the sensory ones.^15^ Thus, images of the nerve roots might reveal a different maturation process.

The comparison of the growth dynamic of the CSA found in this study with the data available in the literature for the motor and sensory nerve conduction velocity of the median nerve^2,17,18^ shows some interesting parallels. The nerve conduction velocity is lowest at the time of birth, but this is also when the increase in nerve conduction velocity is highest. The rate of increase is inversely proportional to age, and by the age of 2 years, the nerve conduction velocity reaches about 70% of its final value. This suggests that a very similar logarithmic model can describe the increase in conduction velocity.

From a molecular physiological viewpoint, it is known that the increase in the nerve conduction velocity is due to an increase in the nerve sheaths and the internodal distances. This increase in the thickness of the myelin, the axon diameter and the internodal distance starts during foetal development at Week 15 of gestation and is rapid during the first year of life.^15^ Combining the models describing the CSA and nerve conduction velocity and taking the molecular basis of maturation of nerves into consideration, a close relation is most likely. This relation should be studied in greater depth in the future using a combination of nerve ultrasound and nerve conduction studies. From a clinical point of view, normal ultrasound data of peripheral nerves combined with neurophysiologic examination of the nerve conduction velocity and electromyography of infants or young children are an essential basis for the diagnosis of early onset diseases that present with a loss of axons and therefore diminished myelin sheath, such as spinal muscular atrophy or white matter diseases (e.g., Krabbe’s disease with an enlarged nerve sheath).^19^

To conclude, this study shows a linear logarithmic growth in the CSA of the median nerve during the first months of life. It further demonstrates that the maturation of the nerve can be analyzed by high-resolution ultrasound imaging and provides normative data for the development of the CSA of the median nerve.

## Data Availability

All measurement data is included in the submission.

## Acknowledgement

We thank PD Hannes Gruber (University of Innsbruck) for helping to initiate the project.

We also thank Monika Hottinger, Roman Rechtseiner and Christoph Simm from Canon Medical Systems for support using the imaging system. We would especially like to thank all participants and their parents for taking part in the study.

## Conflicts of interest

This project was partly funded by Canon Medical Systems.

**Abbreviations**
CSA: Cross-sectional area

---

**Supplementary Figure 1:**
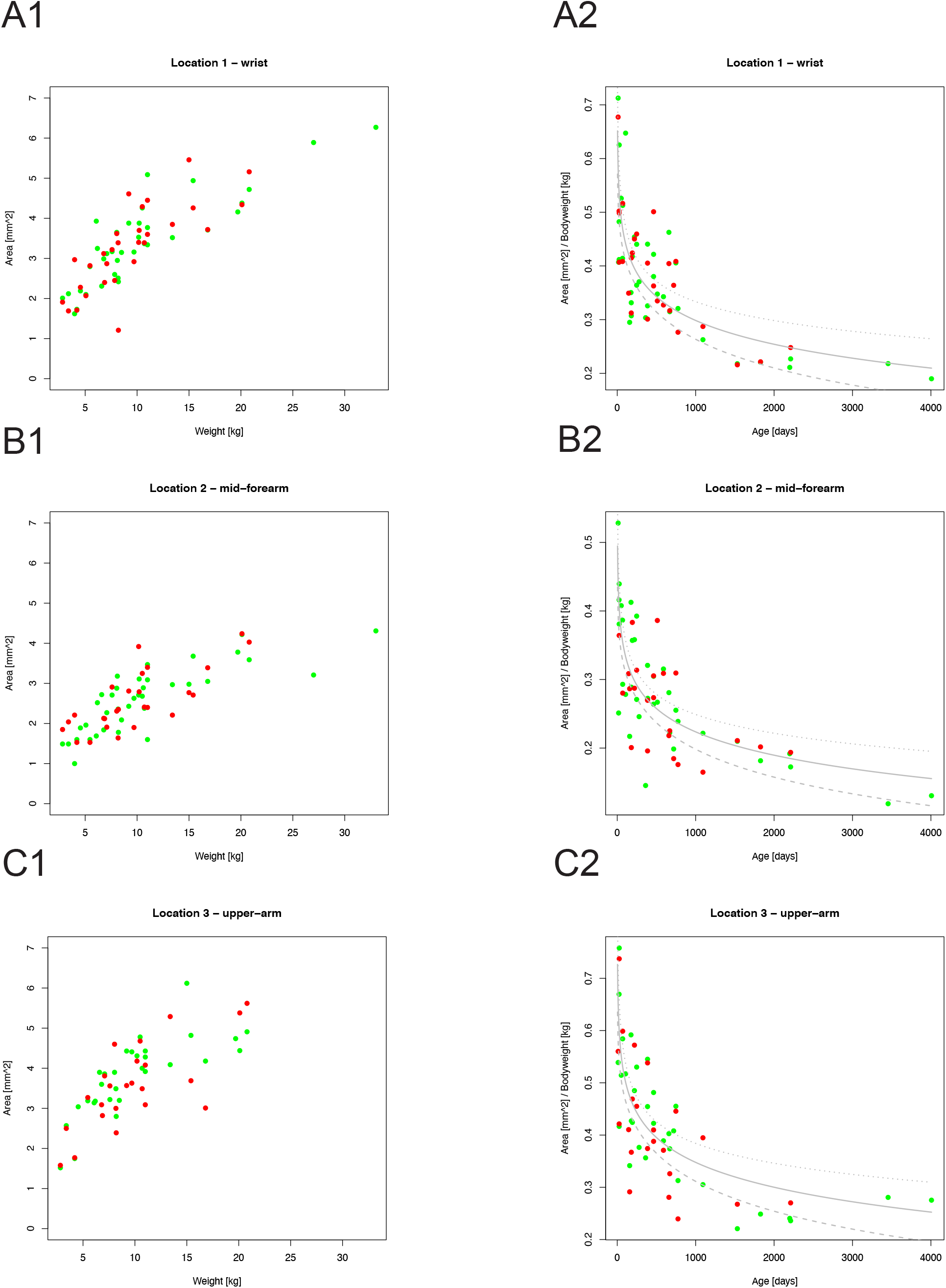
(A.1–C.1) cross-sectional area in relation to weight. Correlation was significant for all three locations. (A.2–C.2) CSA of the median nerve normalized for bodyweight. Normalized CSA followed a logarithmic model with negative slope *a*. Location 1: *a* = −0.063, *b* = 0.740, *p* < 0.0001. Location 2: *a* = −0.049, *b* = 0.563, *p* < 0.0001. Location 3: *a* = −0.069, *b* = 0.822, *p* < 0.0001.

